# Impact of guidance on trends of steroid prescriptions for COVID-19 inpatients: an analysis of the nation-wide administrative database in Japan

**DOI:** 10.1101/2022.12.20.22283717

**Authors:** Takuya Higuchi, Jung-ho Shin, Daisuke Takada, Tetsuji Morishita, Susumu Kunisawa, Yuichi Imanaka

## Abstract

**Background:** Since the outbreak of the coronavirus disease 2019 (COVID-19) pandemic, guidance (“Japanese Guide”) has been published by a working group of several academic societies and announced by the Ministry of Health, Labour, and Welfare. Steroids as a candidate treatment for COVID-19 were noted in the Japanese Guide. However, the prescription details for steroids, and whether the Japanese Guide changed its clinical practice, were unclear. This study aimed to examine the impact of the Japanese Guide on the trends in the prescription of steroids for COVID-19 inpatients in Japan.

**Methods:** We selected our study population using Diagnostic Procedure Combination (DPC) data from hospitals participating in the Quality Indicator/Improvement Project (QIP). The inclusion criteria were patients discharged from hospital between January 2020 and December 2020, who had been diagnosed with COVID-19, and were aged 18 years or older. The epidemiological characteristics of cases and the proportion of steroid prescriptions were described on a weekly basis. The same analysis was performed for subgroups classified by disease severity.

**Results:** The study population comprised 8603 cases (410 severe cases, 2231 moderate II cases, and 5962 moderate I/mild cases). The maximum proportion of cases prescribed with dexamethasone increased remarkably from 2.5% to 35.2% in the study population before and after week 29 (July 2020), when dexamethasone was included in the guidance. These increases were 7.7% to 58.7% in severe cases, 5.0% to 57.2% in moderate II cases, and 1.1% to 19.2% in moderate I/mild cases. Although the proportion of cases prescribed prednisolone and methylprednisolone decreased in moderate II and moderate I/mild cases, it remained high in severe cases.

**Conclusions:** We showed the trends of steroid prescriptions in COVID-19 inpatients. The results showed that guidance can influence drug treatment provided during an emerging infectious disease pandemic.

## Background

Coronavirus Disease 2019 (COVID-19) is an infectious disease caused by Severe Acute Respiratory Syndrome Coronavirus 2 (SARS-CoV-2). The World Health Organization (WHO: World Health Organization) China Office reported a cluster of confirmed pneumonia cases in Wuhan, Hubei Province, China on December 31, 2019. The first COVID-19 case was confirmed in Japan on January 16, 2020. After the Japanese government classified COVID-19 as a designated infectious disease on February 1, 2020, the number of cases increased, and the COVID-19 epidemic has caused tremendous damage to the healthcare system, economy, and society in Japan^1–7^.

Despite the emphasis on evidence-based medicine (EBM), there is a lack of time or human resources to develop typical guidelines under an emerging infectious disease pandemic, but guidance and other information compiled by public agencies on the disease are considered important. EBM is defined as the conscious, explicit, and judicious use of current best evidence in making decisions regarding the treatment of individual patients^8^. In Japan, EBM was introduced in the early 1990s and has since been encouraged by the Ministry of Health, Labour, and Welfare (MHLW). In order for Japanese healthcare professionals to practice EBM, they need knowledge from clinically relevant research articles and reviews^9^. For example, the Medical Information Network Distribution Service, the MHLW-commissioned project to promote EBM, provides support for the development of medical practice guidelines in Japan as well as the evaluation, selection, and publication of such guidelines^10^.

Since the outbreak of the COVID-19 pandemic, many research papers, both clinical and non-clinical, have been published. Governments, international organizations, and academic societies in various countries have published guidance documents on the disease based on this research. In Japan, the “Clinical Management of Patients with COVID-19: a Guide for Front-line Healthcare Workers” (hereinafter, the “Japanese Guide”) was prepared and revised by the Clinical Practice Guidelines Review Committee based on the “Study on the Clinical Response in the Case of Outbreak of Class I Infectious Diseases, etc.” The Japanese Guide has been disseminated by the MHLW’s Novel Coronavirus Infectious Disease Control Promotion Headquarters^11^.

How new guidance can change a physician’s behavior during an emerging infectious disease pandemic was previously reported. For example, the recommendation of the National Health Service in the United Kingdom to change the anticoagulant from warfarin to a direct-acting oral anticoagulant had an effect on prescription^12^. However, the impact of the guidance on prescriptions in Japan has not been clarified. Given the high likelihood of a pandemic similar to COVID-19 occurring in the future and that the probability of experiencing it in one’s lifetime may double in the next few decades^13^, clarifying the impact of the guidance on clinical practice in Japan will help to inform policy decision and discussion of guidance development in similar situations. In addition, an analysis of the prescription patterns of potential therapeutics for COVID-19 inpatients in the University of California COVID Research Dataset (UC CORDS)^14^ has been reported including the daily use percentage of dexamethasone from March 2020 to Jan 2021 in the United States. The epidemiologic characteristics and prescription patterns of COVID-19 inpatients in Japan have been also reported^15^. In that study, patients were classified into three subgroups by oxygen administration and invasive mechanical ventilation/extracorporeal membrane oxygenation, and the proportions of patients administered ciclesonide or a steroid other than ciclesonide during hospitalization in each subgroup were shown. However, time-series trends of the proportions of steroid prescriptions in Japan are not available.

In the present study, we aimed to evaluate the impact of the Japanese Guide on the trends of prescriptions for COVID-19 inpatients in Japan and to contribute to future countermeasures against emerging infectious diseases. Therefore, we focused on steroids, which have been frequently revised in the drugs listed in the Japanese Guide (Supplementary Figure 1). The efficacy of steroids has been reported to vary depending on the severity of the disease^16^, and the fourth edition of the Guide to Clinical Practice, revised on December 4, 2020, strongly recommends the use of steroids for patients with moderate disease II or higher. Therefore, in this study, we classified cases into subgroups according to the severity of disease. In addition to the Japanese Guide and severity of disease, we also considered other guidelines, research articles, drug approvals and supplies, and medical payments that may have affected steroid prescription.

## Methods

### Data Sources and Study Population

Diagnosis Procedure Combination (DPC) data extracted from the Quality Indicator/Improvement Project (QIP) database were used for this study. The DPC/per-diem payment system (DPC/PDPS) is a case-mix patient classification system originally developed in Japan to implement a standardized electronic claims system and provide transparency of hospital performance^17^. The QIP project, administered by the Department of Health Economics and Quality Management, Kyoto University, collects DPC data provided by voluntarily participating acute care hospitals, including both public and private hospitals, all over Japan. The DPC data consist of several files, including the discharge summary (Form 1) and the EF file. Form 1 contains information on medical records including hospital identifiers, patient demographics, admission and discharge date, diagnoses classified as principal, most- and second-most-resource-intensive, or trigger, comorbidities, and complications. The EF-file contains claims data including the date, amounts, content, and reimbursement points of medical practices including surgery, tests, pharmaceuticals, and medical materials^18^.

The International Statistical Classification of Diseases and Related Health Problems, Tenth Revision (ICD-10) codes were used to define the study population (Supplementary Table 1). The inclusion criteria for cases were as follows: 1) an ICD-10 code for COVID-19 (until February 2020: B34.2, after March 2020: U07.1; U07.2.) was entered for the main disease, the disease that triggered hospitalization, and the disease that most invested medical resources (excluding suspected disease); 2) patients aged ≥ 18 years, and 3) cases discharged between January 2020 and December 2020. The exclusion criteria for cases were (1) no missing EF file for each month of hospitalization and (2) pregnant woman or unknown pregnancy status.

The severity of disease was defined by claims codes for the medical practice that was considered to be performed on cases presenting with each severity of disease according to the severity classification described in the Japanese Guide (Supplementary Table 1). Severe cases were defined as those who underwent tracheostomy, endotracheal intubation, ventilation, or extracorporeal membrane oxygenation (ECMO); moderate II cases were defined as those who underwent oxygen administration, high-flow therapy, or non-invasive positive pressure ventilation (NPPV); moderate I and mild cases were defined as those not classified as severe cases or moderate II cases.

### The Proportion of Prescription

The proportion of prescription was calculated by dividing the number of cases in which that particular drug was administered during a given period by the number of cases in which one or more drugs were administered during that period.

The Japanese Guide was developed by a working group involving the Japanese Society of Infectious Diseases, the Japanese Respiratory Society, the Japanese Society of Intensive Care Medicine, the Japanese Pediatric Society, the Japanese Society of Obstetrics and Gynecology, and other academic societies. It was supported by MHLW and disseminated as an administrative communication from the MHLW Headquarters for the Promotion of Countermeasures against New-type Coronavirus Infections. In this study, the steroids listed in the Japanese Guide were included in the analysis. Ciclesonide was listed in the first edition of the Japanese Guide published on March 17, 2020. Steroids were added in the second edition published on May 18, 2020. Steroids except ciclesonide were deleted and dexamethasone was added in the 2.2 edition published on July 17, 2020. “Steroids” in the Japanese Guide do not include ciclesonide and dexamethasone. Also, prednisolone and methylprednisolone were identified as steroids that could be prescribed for the treatment of COVID-19 based on efficacy and dosage form. Steroids are strongly recommended for moderate II or severe cases, while steroids should not be used in cases who do not require oxygenation in the Japanese Guide.

Cases prescribed with the drugs in this study were defined using the first 7-digits of the 12-digits NHI drug codes to represent the substance level following the previous research^19^ (Supplementary Table 1). The NHI drug code is a 12-digit code consisting of four digits for the drug class, three digits for the route of administration and substances, one digit for the dosage form, one digit for the standard drug class within the same class, two digits for the brand identifier within the same standard unit, and one digit for the check digit. A code is assigned to each drug listed in the National Health Insurance (NHI) drug price standard. In Japan, drugs that are approved under the “Act on Quality, Efficacy, and Safety Assurance of Drugs and Medical Devices (Pharmaceuticals and Medical Devices Act)” and required for medication and dispensing by the National Health Insurance are listed in the NHI Drug Price Standards managed by MHLW.

## Results

### Extraction of the Study Population

A flowchart representing the process of selecting the study population is shown in Figure 1. The study population consisted of 8603 cases, of which 410 were severe cases, 2231 were moderate II cases, and 5962 were moderate I and mild cases.

**Figure 1.**
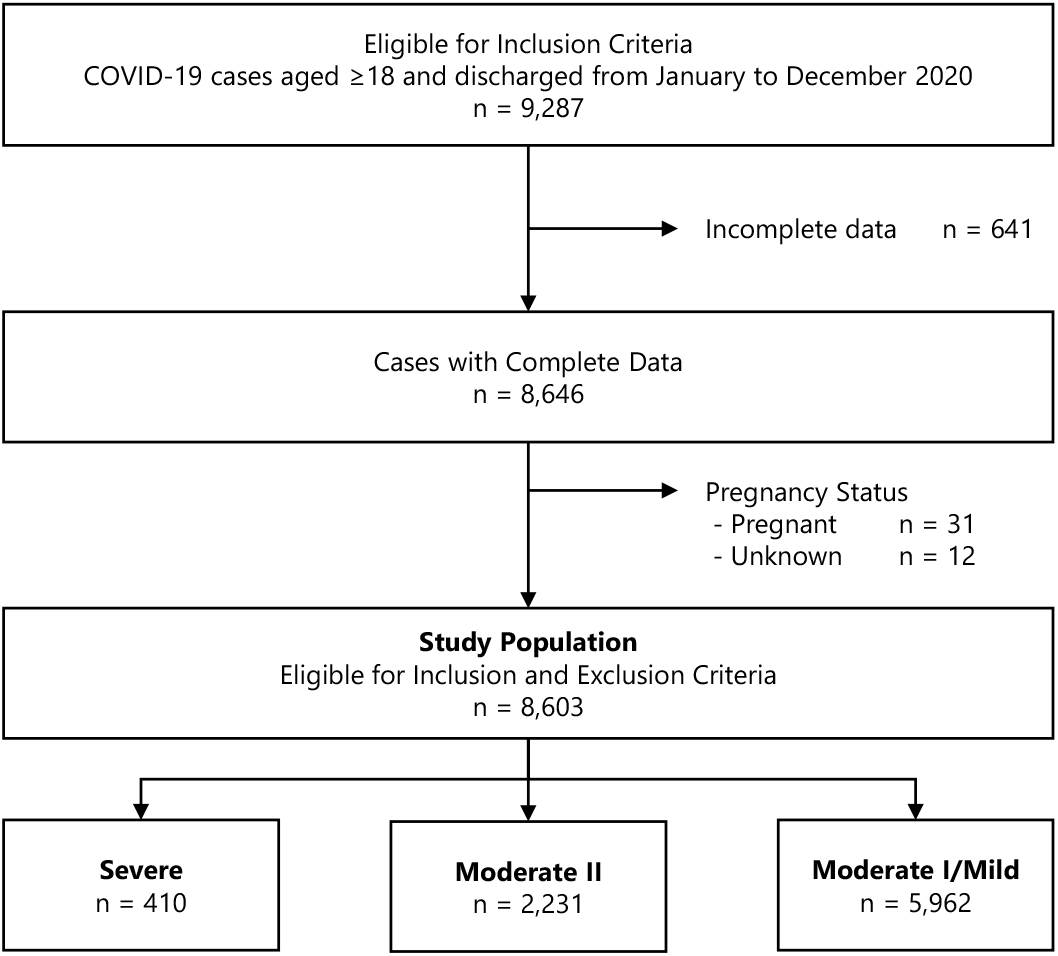
Flow chart of the selection of the study population and subgroups. The number of cases is shown through the selection process.

### Epidemiological Characteristics of COVID-19 Hospitalized Patients

To determine whether the study population and subgroups classified by severity of disease had characteristics consistent with previous reports of COVID-19 patients, their epidemiological characteristics are presented in Table 1. All subgroups had a high proportion of males, and this was particularly evident in severe cases. The median age was higher in severe and moderate II cases than in moderate I and mild cases. The proportion of cases with a history of smoking was lower in severe cases; however, a higher proportion of severe cases had an unknown smoking history. Furthermore, length of hospital stay and in-hospital mortality rates increased with increasing severity of disease.

**Table 1.**
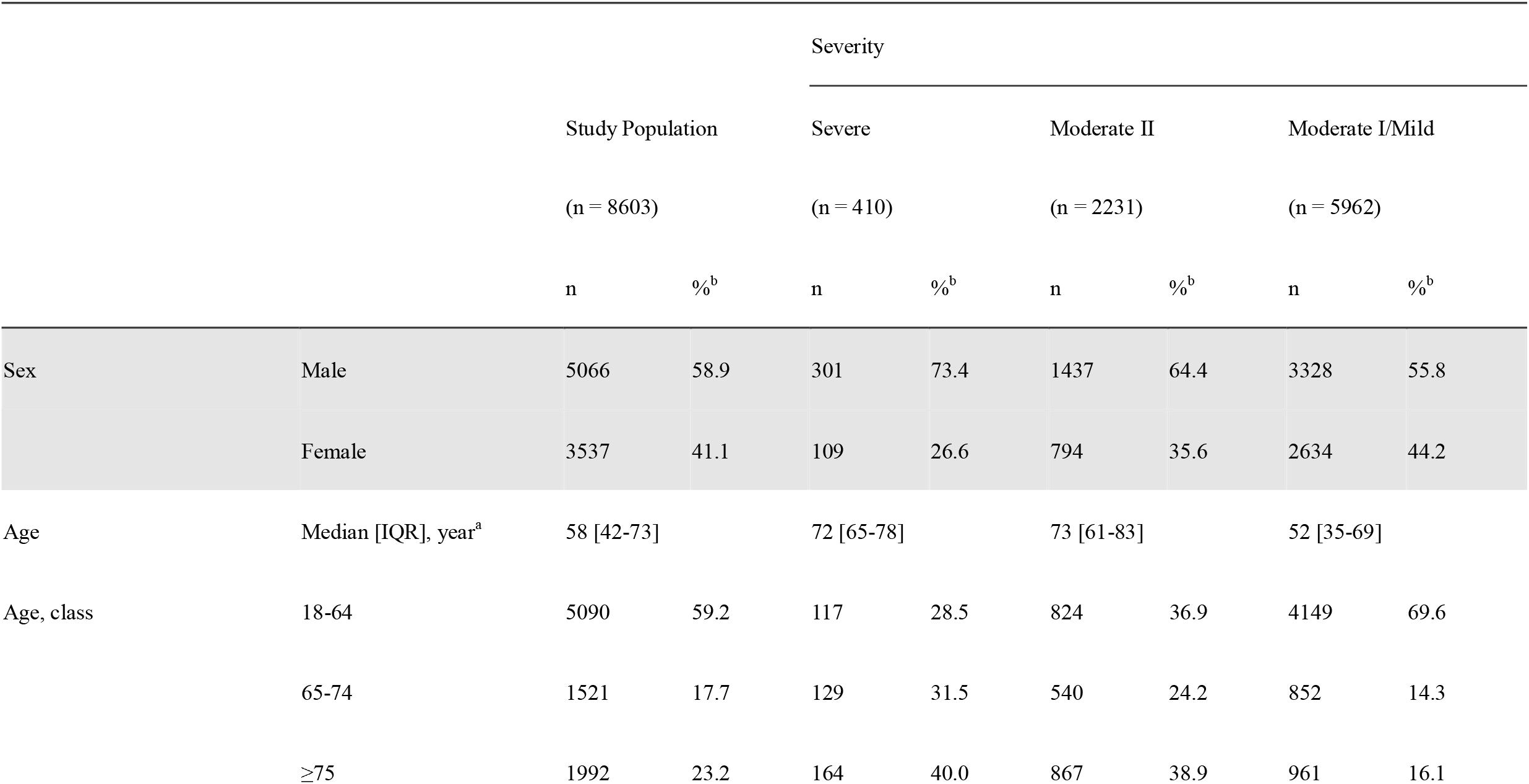
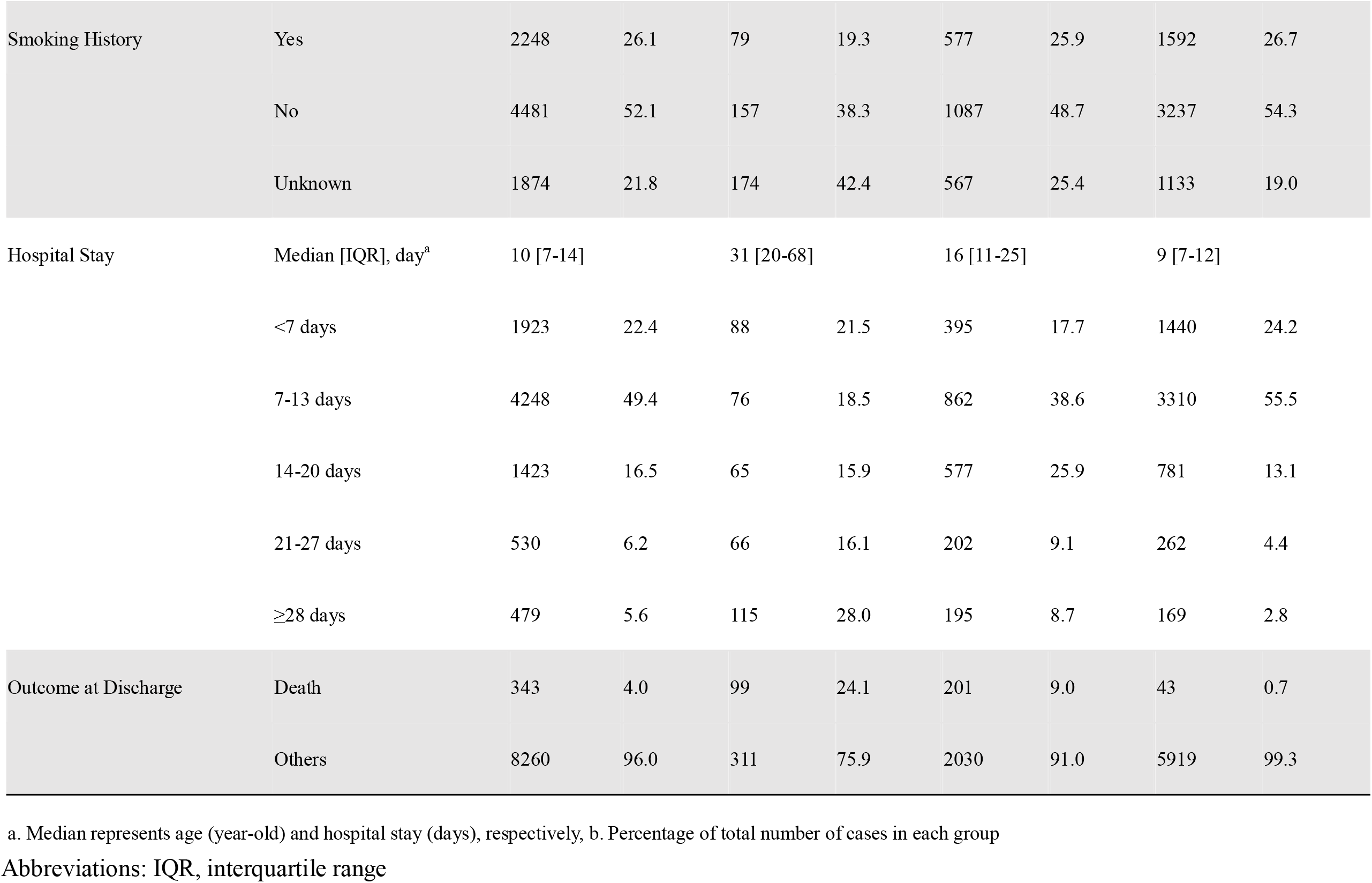
Patient demographics and epidemiological characteristics. Abbreviation: IQR, interquartile range. a. ‘Median’ represents age (years-old) or hospital stay (days), b. Percentage of the total number of cases in each group.

### Number of Cases by Severity of Disease

The number of cases by severity is shown in Figure 2. The increase in the number of hospitalized cases was consistent with the increase in the number of people tested positive for SARS-CoV-2 in April, August, and November 2020, in Japan^20^. Although the proportion of severe/moderate II cases decreased around week 29 (July 2020), when dexamethasone was included in the Japanese Guide, the proportion of severe/moderate II cases did not change before or after that time.

**Figure 2.**
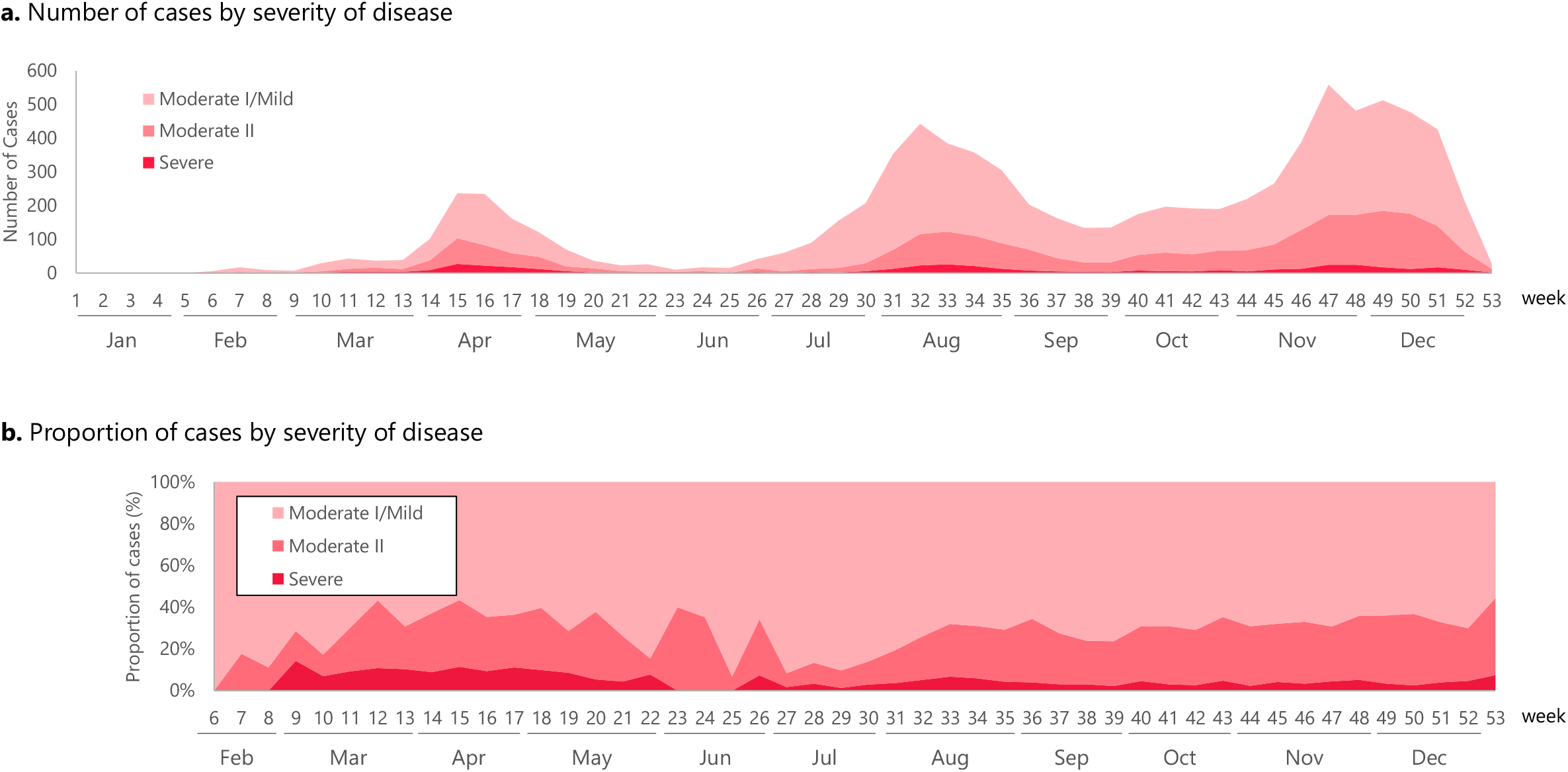
Number and proportion of cases by severity of disease. Figure 2a. Number of cases by severity of disease. Figure 2b. Proportion of cases by severity of disease. (a) Number of cases by severity of disease, (b) Proportion of cases by severity of disease. Horizontal axis represents week number.

### Trends of Steroid Prescriptions

Trends of steroid prescriptions are shown in Figure 3. As shown in Figure 3a, in the study population as a whole, the proportion of dexamethasone prescriptions was less than 5% from January to July 2020 but increased rapidly after July 2020. The proportion of ciclesonide prescriptions reached 17.6% at Week 13 (March 2020) but declined thereafter. The highest proportion of prescriptions was 13.2% at Week 23 (June 2020) for prednisolone and 9.4% at Week 17 (April 2020) for methylprednisolone.

**Figure 3.**
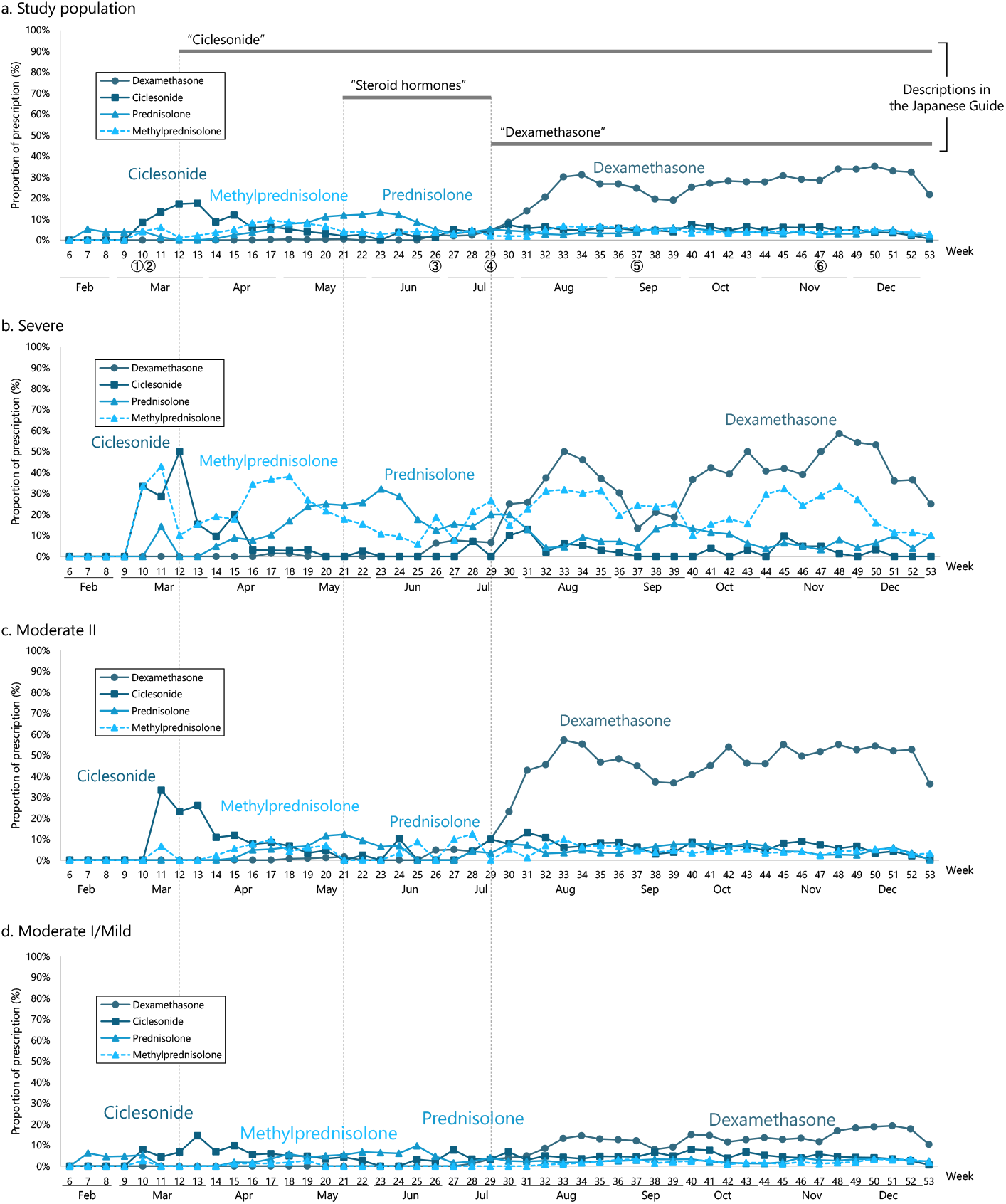
Trends of steroid prescriptions. Figure 3a. Study population. Figure 3b. Severe. Figure 3c. Moderate II. Figure 3d. Moderate I/mild. ① A case report of ciclesonide administration published on the Infectious Diseases Society of Japan website (March 2, 2020), ② Designation of ciclesonide as an drug subject to shipment adjustment (March 3, 2020), ③ Publication of the preprint of the Recovery Trial on the efficacy of dexamethasone (June 22, 2020), ④ Publication of the Recovery Trial in peer-reviewed journals (July 17, 2020), ⑤ Publication of the first edition of “Rapid/Living Recommendations on Drug Management for COVID□19” (September 9, 2020), ⑥ Publication of the first edition of “The Japanese Medical Science Federation COVID-19 expert opinion” (November 20, 2020). Vertical axis represents the proportions of prescription, horizontal axis represents week number. From the fourth edition of the Japanese Guide revised on December 4, 2020, it is recommended that steroids not be administered to the patients with moderate I or lower, while dexamethasone is strongly recommended in moderate II or higher.

In the study population as a whole, several steroids such as ciclesonide, prednisolone, and methylprednisolone were prescribed prior to the publication of the Japanese Guide. However, prescription patterns stabilized after the publication and revision of the Japanese Guide, and dexamethasone became the main treatment option especially after it was included in the guidance. The proportions of prescriptions for steroids other than those listed above were lower than for ciclesonide, prednisolone, methylprednisolone, and dexamethasone.

## Discussion

We analyzed nationwide administrative data to examine the impact of the Japanese Guide on the trends of steroid prescriptions for COVID-19 inpatients. The proportion of dexamethasone prescriptions was remarkably increased after July 2020 when dexamethasone was included in the guidance. Until that update, the Japanese Guide had mentioned “steroid hormones” without specifying a type.

In the past, data on the number of severe cases were reported based on each prefecture’s own criteria. However, in this study, the proportions of severe, moderate II, moderate I, and mild cases among hospitalized cases were clarified in general accordance with the criteria described in the Japanese Guide. In addition, we identified changes in steroid prescriptions for COVID-19 hospitalized cases as well as changes in steroid prescriptions in accordance with continuous revisions of the Japanese Guide. The results suggest that a variety of steroids were prescribed in the early stages of the pandemic, that prescription patterns were not stable, and that the Japanese Guide had some influence on steroid prescription for COVID-19 hospitalized cases.

We analyzed patients’ demographics and epidemiological characteristics to discuss whether the selected study population was appropriate for the aims of the study. As shown in Table 1, there was a high proportion of males in all severity subgroups, and this was particularly pronounced in severe cases. In addition, the median age was higher in severe and moderate II cases than in moderate I and mild cases. It was previously reported that being male^21,22^ and being older than 65 years^23^ were prognostic factors of COVID-19. Furthermore, length of hospital stay and in-hospital mortality rate increased with the rise of severity of disease. In our study, the epidemiological characteristics of each severity subgroup were consistent with these previous reports, and the definition of severity used in this study was considered adequate to classify the patient population. Contrary to our expectations, the proportion of cases with a smoking history was lower in the severe cases, whereas the proportion of cases with an unknown smoking history was higher in the severe cases. This suggests that it may be difficult to obtain a smoking history in severe cases.

As shown in Figure 3, all severity subgroups had a relatively high proportion of prednisolone and methylprednisolone prescriptions in early 2020, but the proportion of dexamethasone prescriptions increased markedly after it was included in the Japanese Guide. The proportions of prescriptions for prednisolone and methylprednisolone decreased in moderate II cases and in moderate I and mild cases but remained high in severe cases. The proportion of cases prescribed steroids in severe cases varied widely from week to week, which may be attributed to the small number of severe cases (410 cases). The proportion of cases prescribed steroids in moderate I and mild cases was lower than in severe and moderate II cases; however, the proportion of dexamethasone prescriptions increased after its inclusion in the Japanese Guide. Furthermore, despite the recommendation not to administer steroids to patients with moderate disease I and mild disease in the fourth edition of the Japanese Guide revised on December 4, 2020, the proportion of dexamethasone prescriptions remained around 20% after the revision. This result suggests that dexamethasone may have been administered regardless of the disease severity.

The Subcommittee on Infectious Diseases of the Health Sciences Council held on December 17, 2020 recommended that, as one of the main issues in the response against the pandemic, it was necessary to consider strengthening research on infectious diseases including collaboration between the National Institute of Infectious Diseases, the National Center for Global Health and Medical Research, and other related organizations. However, this study suggested that the recommendations in the Japanese Guide were not always followed adequately. Therefore, measures should be taken to promote compliance with the guidance, such as using tables and figures to communicate differences in the recommendations of drugs for each severity.

### Factors Other Than the Japanese Guide

In order to assess the impact of the Japanese Guide on steroid prescriptions, it is necessary to consider factors other than the Japanese Guide that may have influenced steroid prescription. In addition to severity of disease, such factors include guidelines, research articles, drug approval and supply, and payments including cost-sharing and reimbursement. The increase in the proportion of dexamethasone prescriptions was not likely due to changes in patient severity because the proportion of severe/moderate II cases did not change before or after Week 29 (July 2020) as shown in Figure 2.

First editions of the “Rapid/Living Recommendations on Drug Management for COVID□19^24^” and the “The Japanese Medical Science Federation COVID-19 expert opinion ^25^”, which are also the major guidelines and guidance in Japan, were published on September 9, 2020 and November 20, 2020, respectively (Figure 2 ⑤, ⑥). Therefore, they could not have caused the increase in the proportion of dexamethasone prescriptions in July 2020. In addition, although ciclesonide attracted attention due to a case report^26^ posted on March 2, 2020 on the website of the Japanese Society of Infectious Diseases, the rapid increase in its demand on the following day (March 3) led to a shipment adjustment, which may have suppressed the increase in the proportion of prescription (Figure 2 ①, ②). One of the reasons for the increase in prescriptions of prednisolone and methylprednisolone after April may have been because they are alternatives to ciclesonide, which was subject to shipment adjustments. However, because the proportion of dexamethasone prescriptions in the study population as a whole remained high despite a temporary drop during a shipment adjustment implemented around September 2020, the impact of the shipment adjustment on the proportion of dexamethasone prescriptions may not have been significant. However, it is difficult to discuss the impact of the Japanese Guide on changes in the proportion of dexamethasone prescriptions. This is because a report on the efficacy of dexamethasone^16^ (following its publication on the preprint server on June 22, 2020^27^) was released on the same day as its inclusion in the Japanese Guide (Figure 2 ④). However, there was no significant increase in the proportion of dexamethasone prescriptions immediately after the publication of the preprint despite the media coverage that preceded the preprint being a hot topic in Japan, including coverage in a national newspaper^28^. Furthermore, causal inferences cannot be performed even when using quasi-experimental designs such as interrupted time-series analysis.

Regarding drug approval, steroids such as dexamethasone were indicated for the treatment of severe infectious diseases, but none were indicated for the treatment of novel coronavirus infection. Thus, no impact was expected on the proportion of prescriptions due to the switch from steroids with no indication of “novel coronavirus infection” to steroids indicated for “novel coronavirus infection”. Finally, no statements were found from the MHLW notice regarding the handling of medical payments that would lead to a change in the proportion of prescriptions of a particular steroid.

### Limitations of the Study

The study had three limitations. First, if factors other than the Japanese Guide influenced steroid prescriptions at the same time, we could not distinguish between the influences of these factors. However, taking these factors into account as described above, it is considered that the Japanese Guide had some influence on Japanese physicians’ decisions regarding prescription during the COVID-19 pandemic. Second, the data from hospitals participating in the QIP project utilized in this study may not be generalizable to the whole of Japan. Because the data utilized in this study were collected from hospitals that voluntarily participated in the QIP project, which aims to evaluate and improve the quality of medical care, they may have had a higher awareness of compliance with guidelines, etc. Thus, we may have overestimated changes in prescription in response to the Japanese Guide. However, because acute care hospitals of various sizes from all over Japan voluntarily participate in the QIP project, bias due to the region or size of the medical institution’s location was considered to be small. Third, there was a possibility of misclassification of disease severity. In this study, severity of disease was defined based on the medical practices included in the DPC data. However, because severity of disease is supposed to be determined objectively by evaluating oxygen saturation, which is not included in the DPC data, it is possible that differences in classifications may have occurred. However, based on the epidemiological characteristics of each severity subgroup shown in Table 1, the differences were considered to be small.

## Conclusions

The present study suggests that the Japanese Guide influenced Japanese physicians’ decisions regarding prescriptions during the COVID-19 pandemic. However, contrary to the recommendations in the Japanese Guide for drug treatment according to severity, steroids were prescribed even for moderate I and mild cases. This was despite the recommendation not to administer steroids since the fourth edition of the Japanese Guide of December 4, 2020. It is possible that the recommendations in the Japanese Guide were not properly communicated.

## Supporting information

Reporting Guideline Checklist

## Data Availability

The information provided to waive consent from study subjects includes the list of data users. The datasets generated during and/or analysed during the current study are available from the corresponding author on reasonable request. The other contact points are Office of Research Promotion, General Affairs and Planning Division, Kyoto University (; Tel: +81-75-753-9301) and the Ethics Committee, Graduate School of Medicine, Kyoto University.

## Acknowledgements

We gratefully acknowledge the participating hospitals in the QIP and their staff.

## Authors’ contributions

TH conceptualized the study, selected methods, analyzed and interpreted the data, wrote the original draft, reviewed and edited the manuscript, and visualized data. TH was a major contributor in writing the manuscript.

JS conceptualized the study, interpreted the data, developed the programming codes for analyzing data, validated and curated the data, reviewed and edited the manuscript, and contributed the funding acquisition.

DT conceptualized the study, interpreted the data, developed the programming codes for analyzing data, validated and curated the data, and reviewed and edited the manuscript.

TM conceptualized the study, interpreted the data, developed the programming codes for analyzing data, validated and curated the data, and reviewed and edited the manuscript.

SK conceptualized the study, interpreted the data, validated and curated the data, and reviewed and edited the manuscript.

YI is the guarantor of the article, and YI conceptualized the study, interpreted the data, validated and curated the data, and reviewed and edited the manuscript. YI supervised and administrated the research project, and contributed the funding acquisition.

All authors read and approved the final manuscript.

## Ethics approval and consent to participate

The authors conducted the present study in accordance with the “Ethical Guidelines for Life Science and Medical Research Involving Human Subjects” issued by the Ministry of Education, Culture, Sports, Science, and Technology, the Ministry of Health, Labor, and Welfare, and the Ministry of Economy, Trade, and Industry. The present study was approved by the Ethics Committee, Kyoto University Graduate School and Faculty of Medicine (approval number: R0135).

## Consent for publication

Consent from study subjects was waived by providing required information to study subjects according to the Ethical Guidelines for Medical and Health Research Involving Human Subjects of the Ministry of Health, Labour and Welfare, Japan (a provisional translation is available from https://www.mhlw.go.jp/file/06-Seisakujouhou-10600000-Daijinkanboukouseikagakuka/0000080278.pdf).

---

**Supplementary Figure 1.**
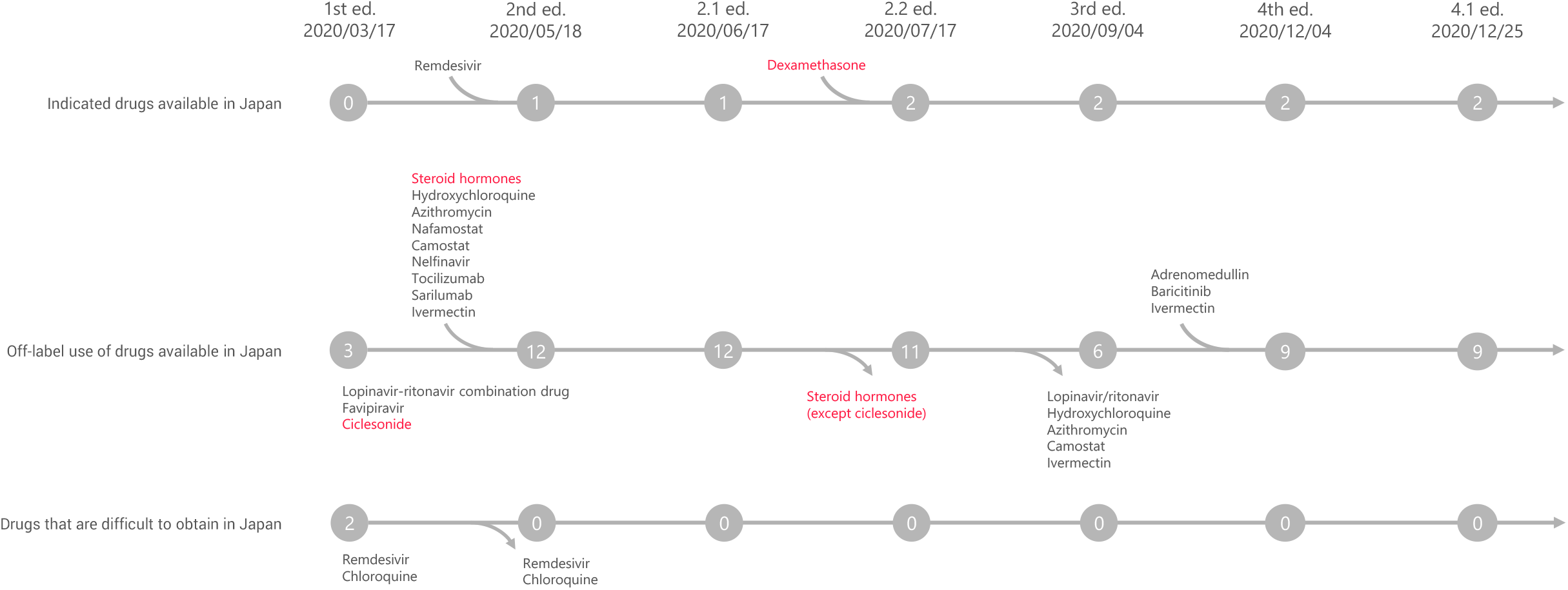
The transition of pharmaceuticals mentioned in “Clinical management of patients with COVID-19: a guide for front-line healthcare workers” The editions of the Japanese Guide and dates of their publication were described. The Japanese Guide have three sections: Indicated drugs available in Japan; Off-label use of drugs available in Japan; Drugs that are difficult to obtain in Japan. The curved lines, joining the horizontal straight lines, mean pharmaceuticals which was added to the Japanese Guide at each revision. The curved arrows, branching out from the horizontal straight lines, mean pharmaceuticals which was deleted from the Japanese Guide at each revision. Numbers in circles are numbers of pharmaceuticals which was mentioned in the Japanese Guide at each edition.

**Supplementary Table 1.**
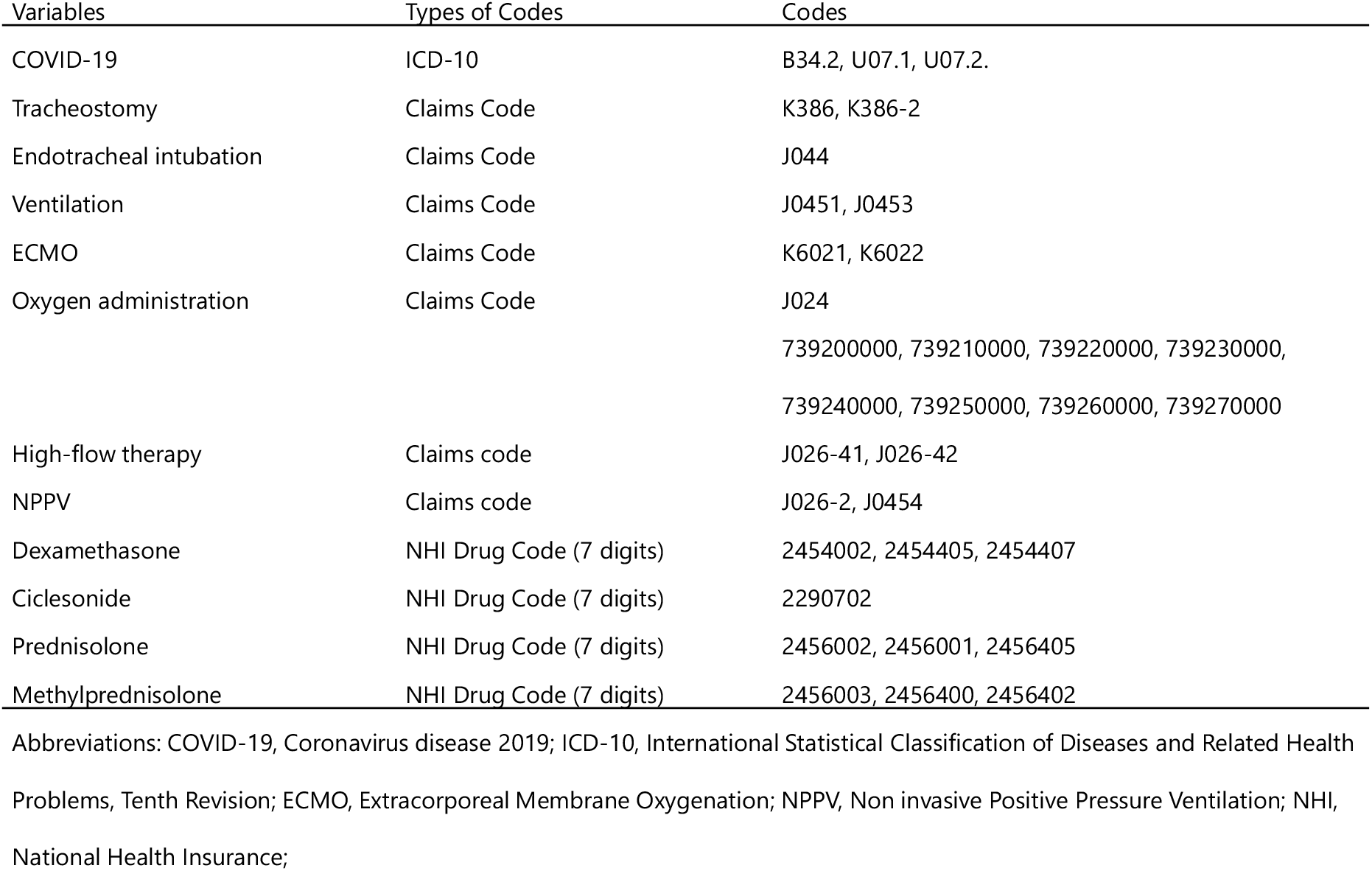
Variables, Types of Codes, and Corresponding Codes

